# Impact of vaccination on the COVID-19 pandemic: Evidence from U.S. states

**DOI:** 10.1101/2021.05.08.21256892

**Authors:** Xiao Chen, Hanwei Huang, Jiandong Ju, Ruoyan Sun, Jialiang Zhang

**Affiliations:** School of International Trade and Economics, University of International Business and Economics; Department of Economics and Finance, City University of Hong Kong; PBC School of Finance, Tsinghua University; Department of Health Care Organization and Policy, School of Public Health, University of Alabama at Birmingham

## Abstract

Governments worldwide are implementing mass vaccination programs in an effort to end the novel coronavirus (COVID-19) pandemic. Although the approved vaccines exhibited high efficacies in randomized controlled trials^1,2^, their population effectiveness in the real world remains less clear, thus casting uncertainty over the prospects for herd immunity. In this study, we evaluated the effectiveness of the COVID-19 vaccination program and predicted the path to herd immunity in the U.S. Using data from 12 October 2020 to 7 March 2021, we estimated that vaccination reduced the total number of new cases by 4.4 million (from 33.0 to 28.6 million), prevented approximately 0.12 million hospitalizations (from 0.89 to 0.78 million), and decreased the population infection rate by 1.34 percentage points (from 10.10% to 8.76%). We then built a Susceptible-Infected-Recovered (SIR) model with vaccination to predict herd immunity. Our model predicts that if the average vaccination pace between January and early March 2021 (2.08 doses per 100 people per week) is maintained, the U.S. can achieve herd immunity by the last week of July 2021, with a cumulative vaccination coverage of 60.2%. Herd immunity could be achieved earlier with a faster vaccination pace, lower vaccine hesitancy, or higher vaccine effectiveness. These findings improve our understanding of the impact of COVID-19 vaccines and can inform future public health policies regarding vaccination, especially in countries with ongoing vaccination programs.

## MAIN

The novel coronavirus (COVID-19) pandemic has had a devastating impact on health and well-being, with more than 131 million cases and 2.8 million deaths across more than 200 countries^3^ as of early April 2021. Despite various regional and national non-pharmaceutical interventions^4-6^ such as travel restrictions, social distancing measures, stay-at-home orders, and lockdowns, many countries continue to struggle with the growth of COVID-19 cases. It is obvious that a successful COVID-19 vaccination program is needed to end the pandemic and allow a return to normal life^7,8^.

By the end of February 2021, two COVID-19 vaccines had been approved in the U.S.: BNT162b2 (Pfizer/BioNTech) and mRNA-1273 (Moderna)^9^. In two large randomized controlled trials (RCTs), the Pfizer vaccine exhibited an efficacy of 95% (95% confidence interval [CI], 90.3% - 97.6%)^1^ in preventing COVID-19, and the Moderna vaccine showed an efficacy of 94.1% (95% CI, 89.3%-96.8%)^2^. Both are mRNA vaccines that require two doses to complete vaccination and received emergency use authorization by the U.S. Food and Drug Administration in December 2020^10^. Mass vaccination campaigns with these two vaccines have since begun. By early March 2021, more than 121 million doses had been administered across the U.S., with over 43 million individuals (∼13% of the population) fully vaccinated with two doses^11^.

Although the efficacies of these two vaccines were shown to be high in RCTs, there is limited information on their potential population-level impact on the COVID-19 pandemic. One peer-reviewed study that estimated vaccine effectiveness used data from nationwide mass vaccination in Israel and reported the effectiveness of the Pfizer vaccine to be 46% (95% CI, 40%-51%) after the first dose and 92% (95% CI, 88%-95%) after the second dose for documented infection^12^. Another study that examined the effectiveness of the Pfizer vaccine among U.S. residents in skilled nursing facilities reported an estimation of 63% (95% CI, 33%-79%) after the first dose^13^.

In this study, we employed well-established reduced-form econometric techniques^14^, commonly used to evaluate the effects of policies or events^15,16^, to assess the impact of vaccination during the ongoing outbreak using data from all 50 U.S. states and the District of Columbia (DC). Although the allocation of vaccines is roughly proportional to state population (Extended Data Fig. 1a), the actual proportion of the vaccinated population differs significantly across states over time (Extended Data Fig. 1b), which provides the key variation to identify the impact of vaccination. Effectively, the observations from each region in the weeks before the vaccination program served as the “control” for the observations after the vaccination program began (“treatment”), with variations in the vaccination rates leading to changes in the “treatment intensity.” By comparing the outcomes across states before and after the initiation of vaccination programs, we evaluated the impact of vaccination on the COVID-19 pandemic.

### Study Design

We collected state-level daily infection and hospitalization data in the U.S. from 12 October 2020 to 7 March 2021. Fig. 1 shows a timeline of COVID-19 developments during this period, including important events and vaccination timeline. We aggregated the data to a weekly level in our baseline estimation given the observed weekly cycle^17,18^ (see Extended Data Table 3 for results using daily data). The dependent variables used to assess the impact of vaccination on the pandemic are the growth rates of total cases and hospitalizations. Our key independent variables are vaccination rates, including the total number of vaccine doses administered per 100 people (at least one dose) and the total number of second doses administered per 100 people. Without any control variables, Fig. 2 shows the negative correlation between the vaccination rate and the growth rates of total cases and hospitalizations.

**Fig. 1.**
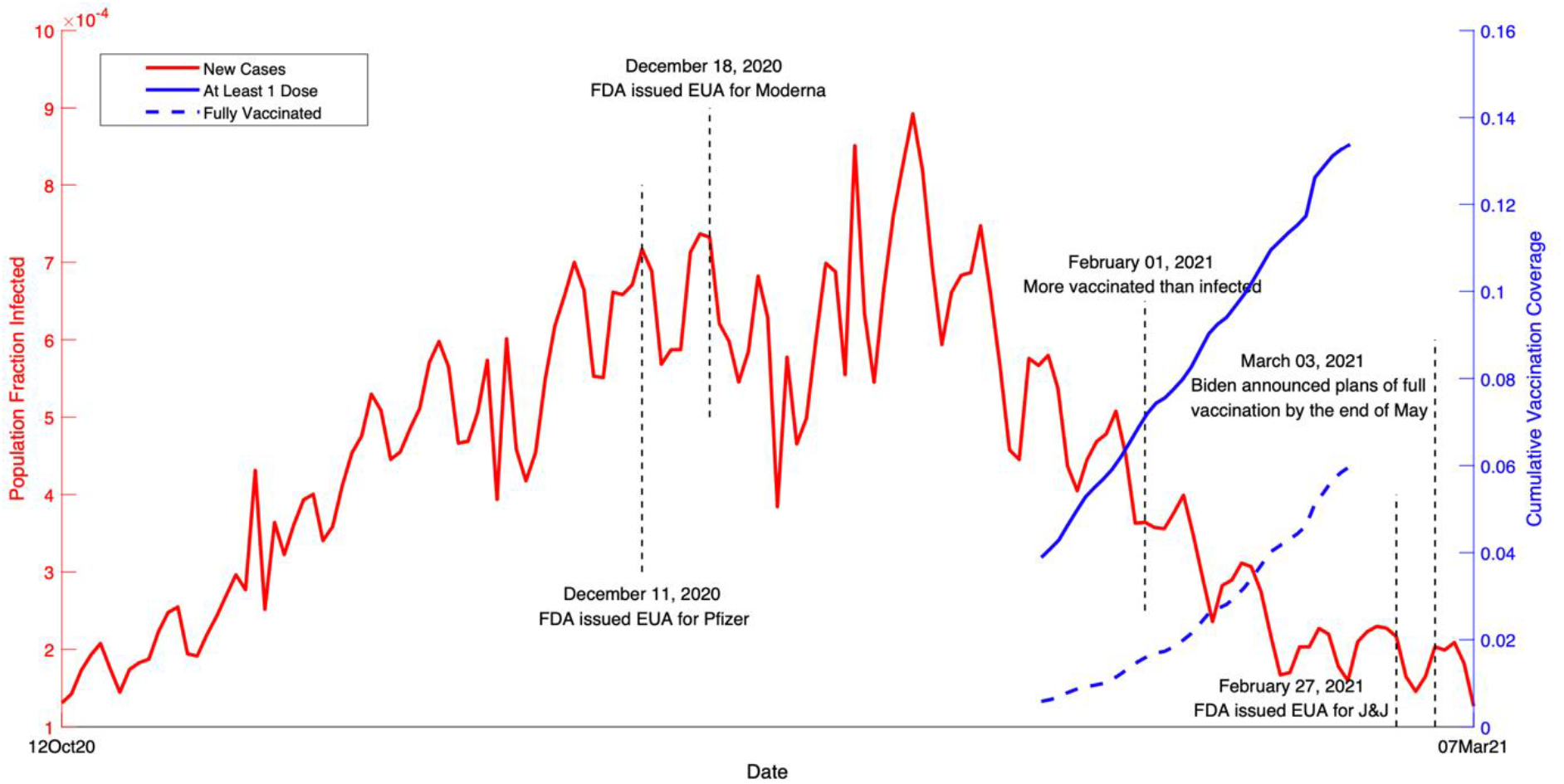
COVID-19 events and vaccination timeline in the U.S. from 12 October 2020 to 7 March 2021. The red curve is the fraction of population infected over time (left y-axis). The solid blue curve is the cumulative vaccination coverage in the population with at least one dose of vaccine (right y-axis). The dashed blue curve is the cumulative vaccination coverage of fully vaccinated individuals in the population (right y-axis).

**Fig. 2.**
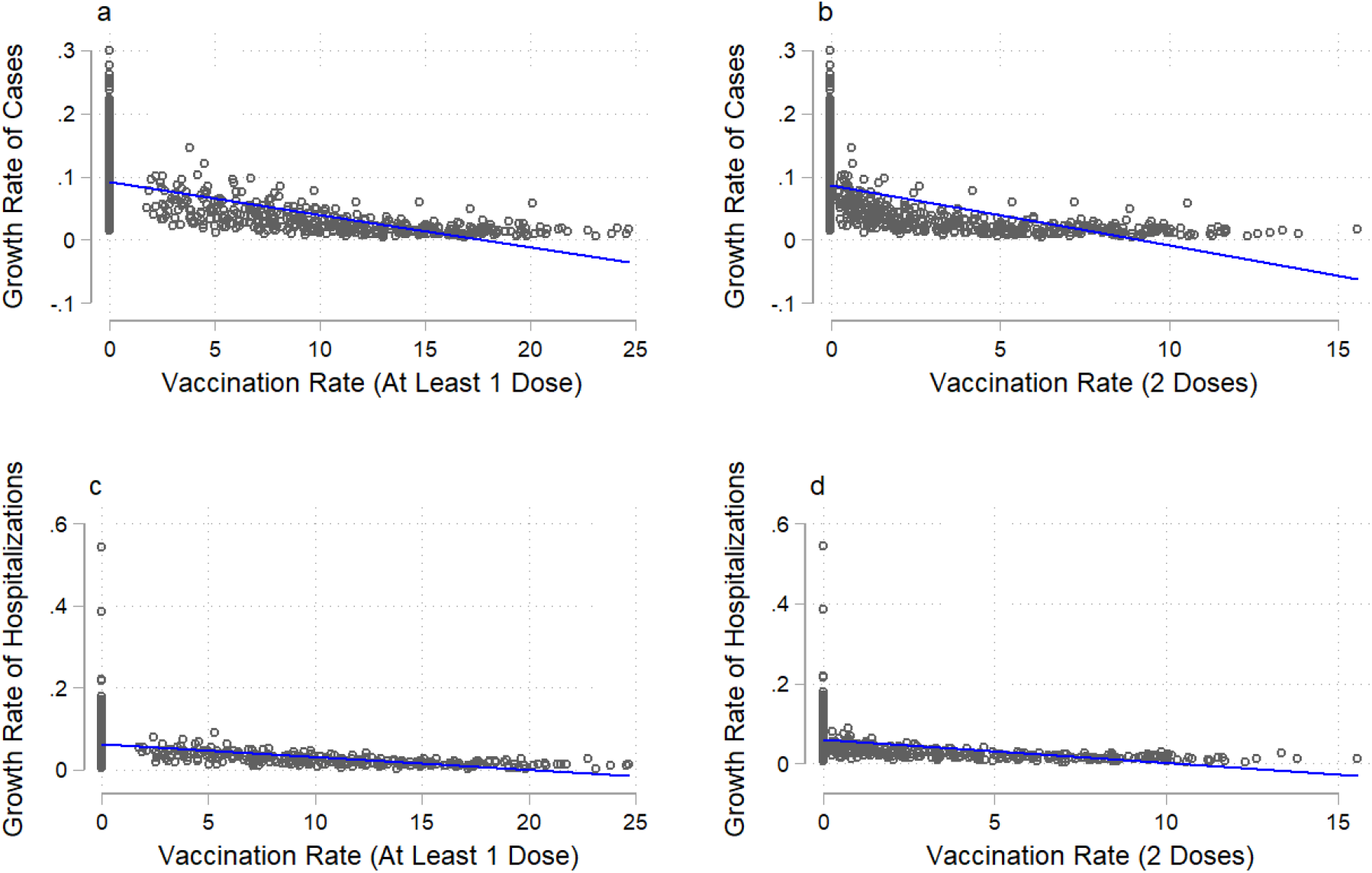
COVID-19 infections (total cases and hospitalizations) and vaccination rate. Vaccination rate is the number of individuals vaccinated per hundred. The solid line in each figure is a fitted linear curve between the growth rate of total cases/hospitalizations and vaccination rate. **a**, Association between the growth rate of total cases and at least 1 dose of vaccination (coefficient = -0.006, R^2^ = 35.3%). **b**, Association between the growth rate of total cases and 2 doses of vaccination (coefficient = -0.013, R^2^ = 28.6%). **c**, Association between the growth rate of total hospitalizations and at least 1 dose of vaccination (coefficient = -0.003, R^2^ = 20.8%). **d**, Association between the growth rate of total hospitalizations and 2 doses of vaccination (coefficient = -0.007, R^2^ = 16.6%).

To make the individual states as comparable as possible, we first accounted for observable factors associated with the COVID-19 pandemic based on previous studies (see Extended Data Table 1). These time-varying control variables included non-pharmaceutical interventions^5-7^, election rallies^19,20^ and anti-racism protests^21^ that involved mass gatherings, and climate measures of snow depth and temperature^22^. To address the concern that changes in the number of total cases reflect the testing capacity of each state^23^, we also controlled for each state’s testing capacity. As the proportion of susceptible individuals declines, the infection rate may slow; therefore, we included the share of susceptible individuals in the regressions. We estimated the dependent variables of COVID-19 cases and hospitalizations with a one-week lag to account for the latency period of infection. Finally, we added state fixed effects and time fixed effects to capture spatial and temporal invariants to alleviate omitted-variable bias.

### Impact of Vaccination

Our data show that the national average weekly growth rate of total cases was 7% (s.e.m. = 0.05) between 12 October 2020 and 7 March 2021. At the individual state level, the average growth rate was highest in Vermont (11%) and lowest in Hawaii (4%). The average growth rate of total hospitalizations across the 35 states that reported hospitalization data was 5% (s.e.m. = 0.04%); the highest growth rate was seen in Montana (8%) and the lowest in New Hampshire (2%).

Vaccination has significantly slowed the growth of total COVID-19 cases and hospitalizations in the U.S. Our baseline results (Fig. 3a and Extended Data Table 2) show that one additional vaccinated individual per 100 people (at least 1 dose) reduced the growth rate of total cases by 0.7% (s.e.m.= 0.2%) and the growth rate of total hospitalizations by 0.7% (s.e.m. = 0.2%). The effects of receiving full vaccination with two doses appear greater, with reductions of 1.1% (s.e.m. = 0.4%) in the growth rate of total cases and 1.1% (s.e.m. = 0.3%) in total hospitalizations. Based on these estimates, vaccination reduced the number of new cases during our study period by 4.4 million (from 33.0 to 28.6 million), which translates into a decrease of 1.34 percentage points in the population infection rate (from 10.10% to 8.76%). Vaccination further reduced the number of hospitalizations by approximately 0.12 million, from 0.89 to 0.78 million (Supplementary Methods).

**Fig. 3.**
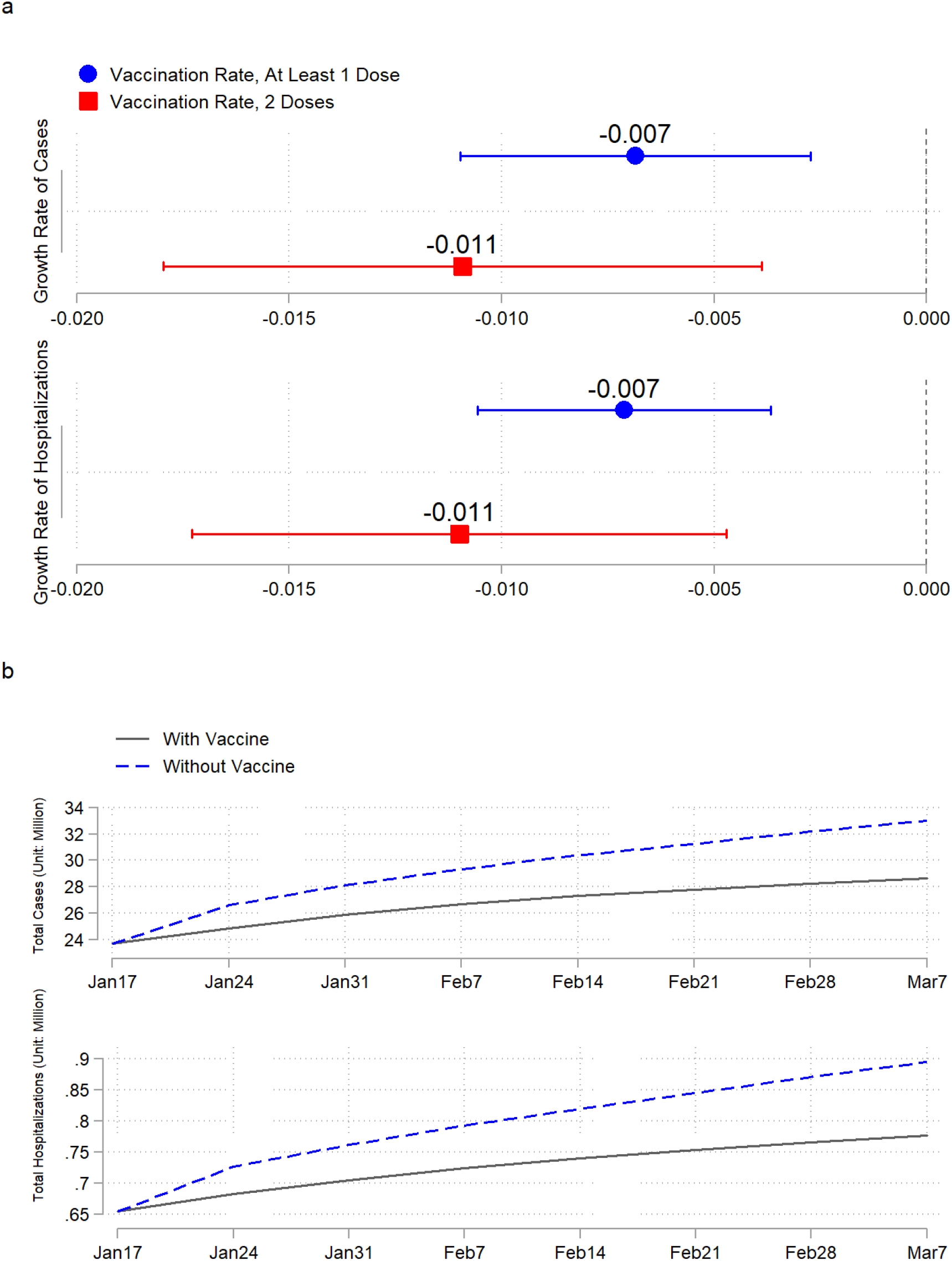
Estimated effects of vaccination on the COVID-19 pandemic. Blue markers are the estimated effects of at least 1 dose of vaccine, and red markers are the estimated effects of 2 doses of vaccine. **a**, Baseline effect of vaccination on the growth rates of total cases and hospitalizations. **b**, Estimated trajectories of total cases and hospitalizations without vaccines (dashed curves) versus actual trajectories of total cases and hospitalizations with vaccines (solid curves).

If systematic correlations existed between the pre-vaccination growth rates of infection and hospitalization and the rate of vaccination, our results would have been subject to selection bias. However, this was not the case. We demonstrated that the number of vaccines allocated to each state was proportional to its population size (Extended Data Fig. 1a). More importantly, we found that the pre-vaccination average growth rates of total cases and hospitalizations were not correlated with the average vaccination rate (Extended Data Fig. 2).

Our baseline results focus on the average treatment effect of vaccination. This effect may be heterogeneous across states that have different characteristics. For example, some evidence shows that the prevalence of COVID-19 differs across age groups, with older adults bearing the highest risk^24,25^. Because older adults were given priority during the rollout of vaccination, it is intuitive to ask whether this strategy made a difference. We separated the states into two groups according to their proportion of older adults (at least 65 years of age). Despite the slightly larger point estimate for the states with a share of older adults above the national median, the results do not differ significantly from those for the states below the median (Extended Data Fig. 3c). In addition to age, we conducted heterogeneity tests on political affiliation, nonpharmaceutical interventions, race, income, and vaccine brand. We found no significant heterogeneous effect of vaccination on any of these characteristics (Extended Data Fig. 3), implying that COVID-19 vaccines have similar effectiveness across these characteristics.

We conducted a range of sensitivity tests. First, instead of using weekly data, we ran regressions with daily data and obtained results of similar magnitudes (Extended Data Table 3). Second, we used alternative measures to capture the development of the pandemic, including the logarithms of new cases and hospitalizations and the changes in logarithms of total cases and hospitalizations. Again, using these measures, we found that vaccination has significantly slowed the pandemic (Extended Data Fig. 4 and Extended Data Table 4). Although the vaccination rollout began on 14 December 2020, our vaccination data begin 11 January 2021; we thus used linear extrapolation to impute the missing data. Our results with the inclusion of imputed data are very similar to the baseline results (Extended Data Fig. 5). Finally, we selected approximately the same number of weeks for the pre-treatment and post-treatment periods to balance the sample in our baseline results. To check the sensitivity of our results to the sample period, we ran our regressions with varying time windows, and our results remain remarkably stable. We obtained approximately the same coefficients for sample periods from 18 to 45 weeks (Extended Data Fig. 6).

### Herd Immunity

To predict how the pandemic will develop with vaccines, and especially when herd immunity might be achieved, we built a Susceptible–Infected–Recovered (SIR) model with vaccination and calibrated it to our data. Our model predictions of the infection rate during the study period showed 99.69% correlation with the empirical data at the national level (Extended Data Fig. 7). Herd immunity is achieved in the model when the real-time basic reproduction number is less than one (Supplementary Methods).

According to our model predictions, at the national average vaccination pace of 2.08 doses per 100 people per week between January and early March of 2021, the U.S. will achieve herd immunity around the last week of July 2021, with a cumulative vaccination coverage rate of 60.2% and a cumulative infection rate of 13.3%. To understand how the speed of vaccination rollout would affect the time needed to reach herd immunity, we simulated herd immunity dates by varying vaccination pace (Fig. 4). We observed a general trend that a faster vaccination pace would allow the U.S. to achieve herd immunity sooner, but with a greater number of total vaccine doses administered and a lower cumulative infection rate. This result can be explained as more individuals gaining immunity from vaccines than from infections if the vaccination pace increases. If the vaccination pace increases to 4 doses per 100 people per week, herd immunity could be reached in early May 2021, but if it decreases to 1 dose, herd immunity would not be achieved until mid-October 2021.

**Fig. 4.**
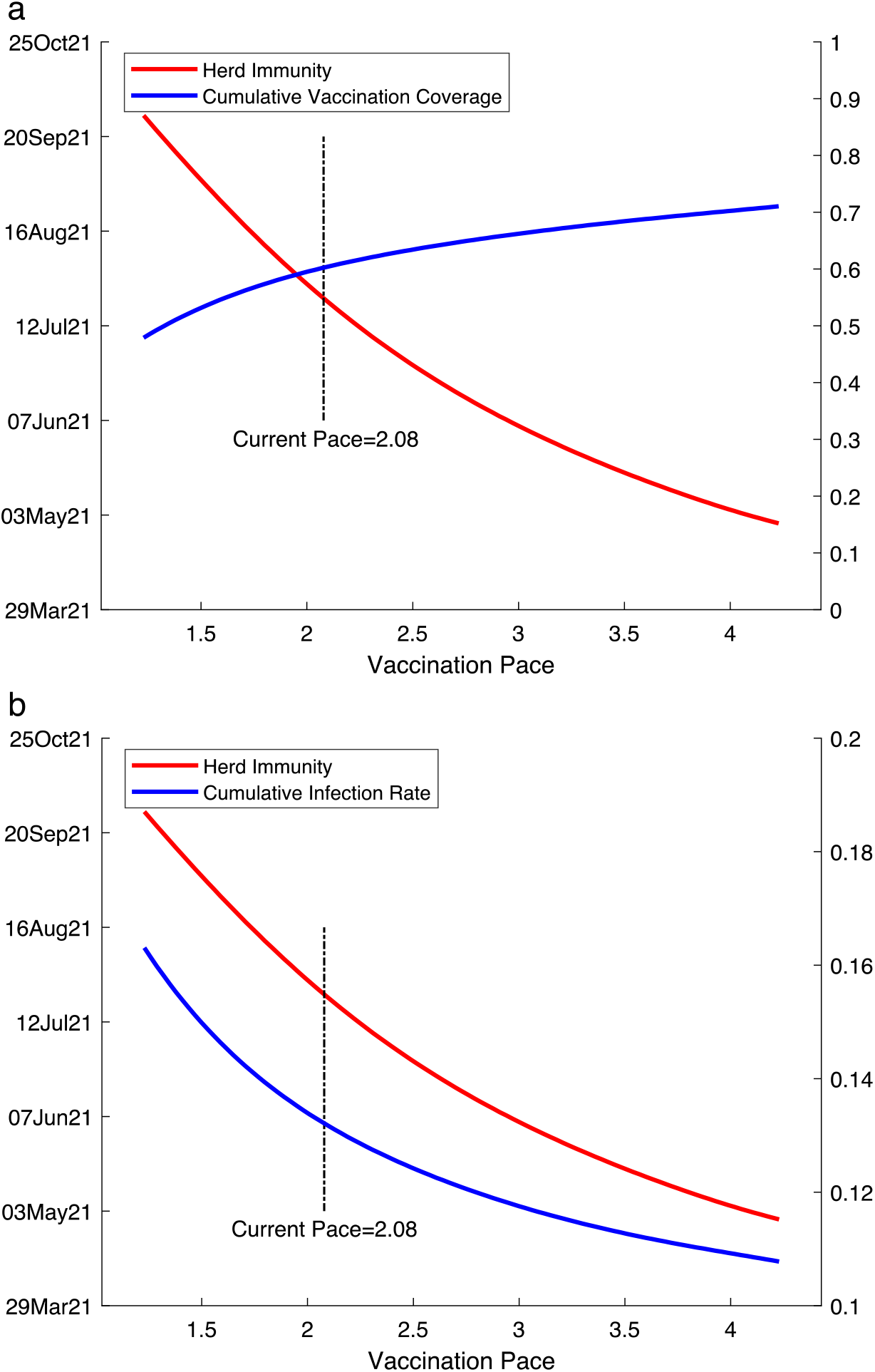
Estimated herd immunity date, cumulative vaccination coverage, and cumulative infection rate with different vaccination pace. Herd immunity date is predicted using first-dose vaccine effectiveness and first-dose vaccination pace (see Methods). Vaccination pace is the number of vaccine doses administered per 100 people per week. Until the first week of March 2021, the average pace over time is 2.08 doses per 100 people per week. The red curve is the predicted herd immunity date (left y-axis) in both Fig. 4**a** and Fig. 4**b**. The blue curve is the estimated cumulative vaccination coverage in the population (right y-axis) when herd immunity is achieved in Fig. 4**a** and the estimated cumulative infection rate (right y-axis) in Fig. 4**b**.

Our predictions of herd immunity assume a continuation of vaccine uptake. In reality, however, a few potential factors could affect uptake. A certain proportion of the population might not receive the vaccination due to vaccine hesitancy. Studies have shown that vaccine hesitancy is a common phenomenon in the U.S.^26,27^, where some individuals are reluctant to receive vaccines due to the perceived risks versus benefits, certain religious beliefs, and a lack of trust in government^27^. Another issue is the effectiveness of vaccines against new coronavirus variants^28^. Only limited evidence is available regarding the efficacy or effectiveness of the Pfizer and Moderna vaccines against these new variants^28^.

To examine how vaccine hesitancy and changes in vaccine effectiveness could affect our predictions for herd immunity, we incorporated in our model a range of potential vaccine hesitancy and vaccine effectiveness estimates. We assumed that if x% of the population is hesitant, then cumulative vaccination coverage in each state will stop when (1 − x%) of the population is vaccinated. Table 1 shows that a higher percentage of vaccine-hesitant individuals will lead to lower vaccination coverage with more individuals infected with COVID-19 at herd immunity. In particular, if vaccine hesitancy reaches 50%, herd immunity will be delayed until the end of August, with a cumulative infection rate of 14.9%; that is, 14.9% of the total population will have been infected with COVID-19 by then. This level of vaccine hesitancy is plausible, given that approximately 40% of U.S. Marines have declined vaccination^29^. In our baseline model, the effectiveness of the first dose of the vaccine was approximately 73% (Supplementary Methods). If the vaccine effectiveness increases, fewer individuals will require vaccination to reach herd immunity, resulting in fewer cumulative cases. However, if the vaccine effectiveness decreases to 60%, herd immunity will not be reached until the week of August 16, 2021, with a cumulative vaccine coverage of 67.6% and a cumulative infection rate of 14.7%.

**Table 1.** Predicted herd immunity with different vaccination pace, vaccine hesitancy, and vaccine effectiveness estimates.

|  | **Herd Immunity Date<sup>a</sup>** | **Vaccination Coverage<sup>b</sup>** | **Fraction Infected<sup>b</sup>** |
| --- | --- | --- | --- |
| **Pace = 1.5** |  |  |  |
| *Vaccine Hesitancy<sup>c</sup>* |  |  |  |
| 10% | 06 Sep 2021 | 53.6% | 15.2% |
| 30% | 06 Sep 2021 | 53.3% | 15.2% |
| 50% | 06 Sep 2021 | 48.4% | 16.3% |
| *Vaccine Effectiveness<sup>d</sup>* |  |  |  |
| 60% | 27 Sep 2021 | 58.1% | 16.7% |
| 80% | 23 Aug 2021 | 50.6% | 14.6% |
| 100% | 26 Jul 2021 | 44.6% | 13.3% |
| **Pace = 2.08 (national average pace between January and early March in 2021)** |  |  |  |
| *Vaccine Hesitancy* |  |  |  |
| 10% | 26 Jul 2021 | 61.4% | 13.4% |
| 30% | 26 Jul 2021 | 60.9% | 13.9% |
| 50% | 30 Aug 2021 | 50.0% | 14.9% |
| *Vaccine Effectiveness* |  |  |  |
| 60% | 16 Aug 2021 | 67.6% | 14.7% |
| 80% | 12 Jul 2021 | 57.2% | 12.9% |
| 100% | 14 Jun 2021 | 48.9% | 11.8% |
| **Pace = 2.5** |  |  |  |
| *Vaccine Hesitancy* |  |  |  |
| 10% | 28 Jun 2021 | 63.6% | 12.6% |
| 30% | 28 Jun 2021 | 63.1% | 13.2% |
| 50% | 20 Sep 2021 | 50.0% | 14.3% |
| *Vaccine Effectiveness* |  |  |  |
| 60% | 26 Jul 2021 | 73.5% | 13.8% |
| 80% | 14 Jun 2021 | 58.6% | 12.1% |
| 100% | 24 May 2021 | 51.1% | 11.1% |

Notes.
<sup>a</sup> Estimated week when herd immunity is achieved. The date mentioned in each row marks the first day of the week.
<sup>b</sup> Cumulative values when herd immunity is achieved.
<sup>c</sup> Percentage of the population who are hesitant to get the vaccine.
<sup>d</sup> Population-level effectiveness of the first dose of COVID vaccines.

A few unanswered questions could still affect herd immunity. One key issue is how long the vaccine immunity will last. Definitive evidence regarding the duration of immunity protection is lacking^30^. Another issue is moral hazard, that is, whether vaccinated individuals will change their behaviors and undertake more social interaction^31^. This change could result in higher risks of infection and a delay in reaching herd immunity.

Our study has a few limitations. We covered only the early periods of vaccination rollout, when the demand for vaccines was greater than the supply. As more individuals become vaccinated, the vaccination pace will likely slow due to the decrease in demand. In addition, our model predictions assume a continuation of the non-pharmaceutical interventions in place in early March. Relaxation of these policies would likely increase the time needed to reach herd immunity. Our SIR model assumes that only susceptible individuals undergo vaccination. However, in real life, many individuals who recovered from COVID later received vaccines. As a result, our model predictions are optimistic, and herd immunity will be achieved later based on this empirical fact. In addition, our study assessed the effects of vaccination in the U.S. using mRNA vaccines. More studies are needed to study vaccination in other countries using different types of vaccines.

Our study provides strong evidence that vaccination has significantly decreased COVID-19 cases and hospitalizations in the U.S. At the average pace of vaccination between January and early March, our model predicts that herd immunity will be achieved around the last week of July 2021, with a cumulative vaccine coverage of 60.2% and a total infection rate of 13.3%. These findings provide grounds for optimism that the pandemic will end during 2021 in the U.S. However, a few factors, such as moral hazard, vaccine hesitancy, and variants of the SARS-CoV-2 virus, could lead to changes and cast doubt as to whether herd immunity can be achieved after all.

## Data Availability

The datasets and code used for the analyses are available at https://github.com/huntabaobao007/US-COVID-19-vaccination.

## METHODS

### Data Collection and Processing

A summary is provided of the data used in our analysis. Our supplementary notes give further details, including a summary statistics table for all variables.

### Epidemiological and Vaccination Data

We collected our state-level epidemiological data (total COVID-19 cases, hospitalization, and tests) from the COVID Tracking Project^32^, a commonly cited source^33-35^. The vaccination data across states were obtained from the U.S. Centers for Disease Control and Prevention’s (CDC) COVID data tracker^36^, where “people vaccinated” reflects the total number of people who have received at least one vaccine dose, and “people fully vaccinated” reflects the number who have received both doses prescribed by the vaccination protocol. We downloaded the CDC vaccination data from an open-source GitHub project by Our World in Data^37^. Both the BNT162b2 (Pfizer/BioNTech) vaccine and the mRNA-1273 (Moderna) vaccine require two doses^9^. In addition, the CDC shares data on COVID-19 vaccine distribution allocations by state for both the Pfizer^38^ and Moderna^39^ vaccines, as provided by the Office of the Assistant Secretary for Public Affairs under the U.S. Department of Health & Human Services.

### Nonpharmaceutical Interventions

In addition to epidemiological data, we obtained information on nonpharmaceutical intervention policies. We adopted the policy stringency index constructed by the Oxford COVID-19 Government Response Tracker^40^, which systematically collects information on various policy responses implemented by various governments in response to the pandemic. We focused on the policy category of “containment and closure,” which covers eight policies: school closings, workplace closings, cancelation of public events, restrictions on gathering sizes, cessation of public transportation, stay-at-home requirements, restrictions on internal movement, and restrictions on international travel. This stringency index is a weighted score across these eight containment and closure policies and is scaled between 0 and 100. A detailed explanation of these measures was given by Hale et al. (2021)^41^. We determined the stringency index for each state on a weekly basis by averaging the daily data.

### Meteorological Data

Another set of important independent variables included in this study regarded the local climate. We obtained station-level hourly weather data provided by the National Centers for Environmental Information^42^. These station-level weather data were then matched with the station location and corresponding state provided by the Global Historical Climatology Network Daily^43^. We calculated the average values from these weather reports for each week across all stations within each state. Given the lack of humidity data, temperature and snow depth were used as our climate measures.

### Election Rallies and Black Lives Matter (BLM) Demonstrations

Several large-scale mass gatherings for political campaigns and protests also occurred during our study period. We constructed binary measures for election rallies^44^. For states with a rally during week *t*, this binary measure takes the value of 1 for week *t* and for the week after (*t*+1). Our BLM data from Elephrame offered detailed information (date, location, etc.) about each demonstration from news reports^45^, which were extracted using a Web scraper. We then calculated the total number of demonstrations that occurred across all cities within each state for each week.

### Sociodemographic Data

We also collected the sociodemographic characteristics of each state’s population using 2019 estimates from the U.S. Census Bureau^46,47^. Specifically, we downloaded data on age, race, and income. We constructed each of our sociodemographic variables to be binary, above or below the national median. We derived the proportion of individuals 65 years of age and older in the population, the proportion of the white population, and the income for each state to calculate a national median. Finally, our data for the 2020 Electoral College results were obtained from the National Archives^48^. We classified the states into those won by Joe Biden and those by won by Donald Trump.

### Econometric Analysis

#### Reduced-Form Analysis

The following reduced-form empirical model was used to estimate the impact of vaccination on the pandemic:

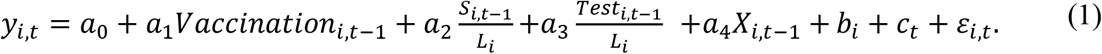

Here, *y*_*it*_ is the dependent variable that measures the growth of either total cases or total hospitalizations in state *i* at period *t*. Our baseline measure is the growth rate, which is defined as 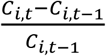for total cases and 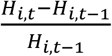 for total hospitalizations, where *C*_*i,t*_ and *H*_*i,t*_ are the cumulative numbers of cases and hospitalizations. Alternative outcome measures were also used in the sensitivity analysis (Extended Data Fig. 4).

Our key independent variable, *Vaccination*_*i,t*−1_, is the rate of vaccination of state *i* in period *t-*1, and *a*_1_ is the coefficient of interest. We used two measures of vaccination rate: the number of vaccinated people (i.e., those who had received at least one dose of vaccine) per hundred and the number of fully vaccinated people (i.e., those who had received two doses of vaccine) per hundred. As the proportion of susceptible individuals in the total population decreases over time, the growth rate of infection may also decline. To deal with this intrinsic dynamic, *S*_*i,t*−1_/*L*_*i*_ was included in the regression model to control for the stock of susceptible individuals *S*_*i,t*−1_ in the total population *L*_*i*_. We measured *S*_*i,t*−1_ as the difference between the population size and the total number of infections. To adjust for differences in testing intensity across states, we added *Test*_*i,t*−1_/*L*_*i*_ to control for the number of tests relative to the total population.

Our control variables contain a dummy variable *rally*_*i,t*_, which equals 1 when an election rally occurred in state *i* at period *t*. We also added a variable *protest*_*i,t*_, which is the number of protests held across all cities in state *i* at period *t*. To capture the influence of climate on the pandemic, we included measures of state-level meteorological conditions, including average temperature, temperature deviation from the state mean, and the logarithm of the average snow depth. Note that we included state fixed effects (*b*_*i*_) to capture state-specific unobserved factors, which are time-invariant, such as location, geography, and culture. We also included week fixed effects (*c*_*t*_) to capture unobserved shocks, which are common across states, such as macroeconomic conditions. Finally, *ε*_*i,t*_ is a random error term of the model, which has a mean of zero.

We estimated equation (1) using the method of Ordinary Least Square with weekly data for 50 states and DC in the baseline. Robust standard errors for the estimated coefficients with two-way clustering were calculated at the state and week levels^49^. Therefore, we allowed for within-state autocorrelation in the error term to capture the persistence of the pandemic within each state. We also allowed for spatial autocorrelation in the error term to capture common pandemic shocks or systematic misreporting across states.

#### Model Summary

We modified a conventional SIR model with the addition of vaccination to simulate the development of the COVID-19 pandemic in the U.S. with vaccine rollout, including both state-level and national-level estimates. The theoretical SIR model with vaccination is as follows:

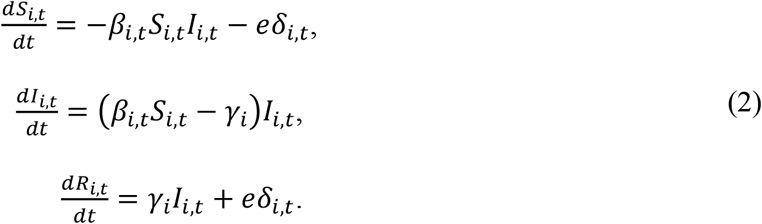

Here, *S*_*i,t*_ is the state-specific (*i*) and time-varying (*t*) proportion of susceptible individuals in the population, *I*_*i,t*_ is the proportion of infected individuals, and *R*_*i,t*_ the proportion of recovered (plus dead) individuals. *β*_*i,t*_ is the infection rate, which determines the spread of the pandemic. *γ*_*i*_ includes both recovered individuals and deaths and is referred to as the removal rate^5^. Here *γ*_*i*_ varies only by state and not over time. *δ*_*i,t*_ is the proportion of vaccinated individuals, and *e* is the population-level vaccine effectiveness, which remains the same across states and time.

We fit the SIR model above with state-level COVID-19 epidemiology data, from which we observed data on the cumulative number of cases, deaths, and vaccination doses administered. Only 29 of the 51 states (counting DC as a “state” for this purpose) reported valid recovery data. We imputed the missing data for the other 22 states with the median recovery and mortality rates from the known 29 states (see Supplementary Methods for details). We first estimated the infection rate (*β*_*i,t*_) and vaccination coverage (*δ*_*i,t*_). To capture the impact of nonpharmaceutical interventions on the spread of COVID-19^4-6^, we used the following equation to estimate the infection rate with state fixed effect (*ρ*_*i*_) and time fixed effect (*ρ*_*t*_):

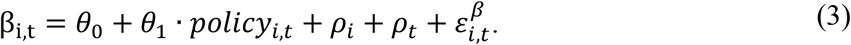

Similarly, we estimated vaccination coverage using the following equations, controlling for state and time fixed effects.

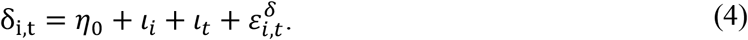

We adopted two vaccination measures in our data: the total number of people who had received at least one vaccine dose and the total number of fully vaccinated people. No time trends were observed in the total doses administered for at least one dose of vaccine, but an apparent time trend was seen in the doses administered for the second dose. We therefore added a time trend in the estimation equation above when we conducted the sensitivity check using the total number of fully vaccinated people as our measure of vaccination. We used equations (3) and (4) to estimate the infection rate and vaccination coverage, combined with the initial epidemiological data of SIR in week 1 (12 October 2020), and our model estimates of the infection rate for the following 20 weeks are highly correlated with the empirical data. For each individual state, our model estimates reached a median correlation of 99.04% (range, 86.37% to 99.95%) (Extended Data Fig. 7).

We assessed herd immunity based on our model estimates of the real-time basic reproduction number for each state, 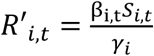 that is, the number of cases directly caused by an infected individual throughout his or her infectious period. The model achieves herd immunity when *R*′_*i,t*_ falls below 1 in 49 states (except for Maryland and Kentucky; see Supplementary Methods for details).

For each given vaccination pace, we ran the simulation forward and projected the future dynamic of the pandemic across the U.S., assuming that no changes are made in nonpharmaceutical interventions. We then computed the time required for every state to achieve herd immunity and calculated the share of the U.S. population vaccinated when herd immunity is achieved. In addition, we conducted a sensitivity analysis regarding herd immunity with variations in vaccine effectiveness and with the addition of vaccine hesitancy. We incorporated vaccine hesitancy into our model by assuming that if x% of the population is hesitant, the cumulative vaccination coverage in each state will stop when (1 − x%) of the population is vaccinated.

## Supplementary Information

The supplementary information provides supplementary methods, figures, and tables.

## 1. Supplementary Methods

### 1.1 Estimating the impact of vaccination

We used our reduced-form estimates to carry out back-of-the-envelope calculations to derive the number of new cases prevented by vaccination. For this purpose, we first calculated the counterfactual growth rate of total cases by *ŷ*_*i,t*_= *y*_*i,t*_−*â*_1_*Vaccine*_*i,t*−1_with *â*_1_and *Vaccine*_*i,t*−1_. *â*_1_ is the estimated effect of vaccination on the growth rate of total cases, and *Vaccine*_*i,t*−1_ is the observed vaccination rate for the previous period. We know that the growth rate *y*_*it*_ satisfies *C*_*i,t*_ = *C*_*i,t*−1_(1 + *y*_*i,t*_), where *C*_*i,t*_ is the observed number of total cases. With the series of counterfactual growth rates of total cases 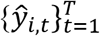 (*T* is the last period of the sample) and a given initial value of total cases (*C*_*i*,0_) before vaccination began, we can infer a counterfactual (without vaccination) series of total cases 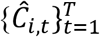 using

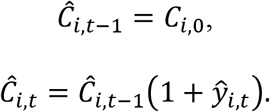

Summing across 50 states and DC, the impact of vaccination on the number of total cases (Δ*C*) is given by

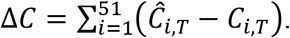

Repeating this process with hospitalization data, we evaluated the impact of vaccination on the total number of hospitalizations during our sample period.

### 1.2 Estimation of Model Parameters

Here we provide more details on parameter estimation for our SIR model with vaccination. We used state-level weekly epidemiological and vaccination data for the estimation during the period from 12 October 2020 to 7 March 2021. The data demonstrate the cumulative population share of infected individuals 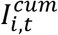 and COVID-19–related deaths 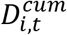 for all 50 states and DC, and valid recovery data 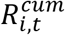 for 29 states^1^. For states with recovery data, we calculated the proportion of infected, susceptible, recovered, and dead individuals for the current period *t* using the following equations: 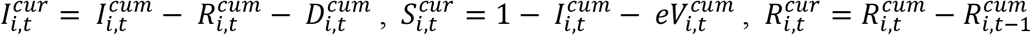 and 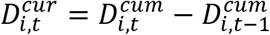, respectively.

### Removal rate (γ_i_)

In equation (2), γ_i_ stands for the removal rate from the infection group. We calculated a state-specific but time-invariant γ_i_ by considering both recovered individuals and deaths following Hsiang et al. (2020)^2^. We obtained complete death data over the study period, but valid recovery data are available for only 29 states. Therefore, we first calculated the average recovery and mortality rates in the 29 states for which we have valid recovery data as

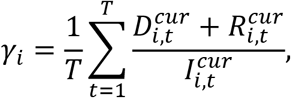

where *T* is the number of weeks in the sample period. The removal rates in the remaining 22 states were assumed to be the median of the removal rates in the 29 states for which complete recovery data are available, that is, 30.15%^3^.

#### Infection rate (β_i,t_)

In our SIR model, β_i,t_determines the spread of the pandemic. According to equation (2), we have

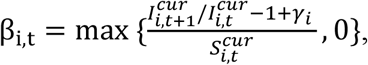

which we used to calculate β_i,t_ in the 29 states that we have recovery data to derive the removal rate *γ*_*i*_ directly. To estimate β_i,t_ for the other 22 states with no recovery data, we first assumed that β_i,t_ is determined by the stringency of nonpharmaceutical interventions and used the following reduced-form equation,

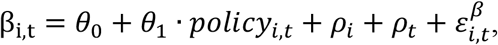

which estimates β_i,t_ for the 29 states with recovery data using the observed nonpharmaceutical interventions, along with state fixed effects (*ρ*_*i*_) and time fixed effects (*ρ*_*t*_). We then inferred β_i,t_ for the remaining 22 states^4^ based on the estimated *θ*_1_, the observed policies, and the median estimates of state and time fixed effects. We also assumed that future non-pharmaceutical interventions would remain at the same level as in the last week of our sample (i.e., the week of March 1, 2021) when generating model predictions.

#### Vaccination rate (δ_i,t_)

We calculated the population share of newly vaccinated people by 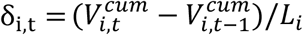, where L_i_ is the total population size in state. We then estimated δ_i,t_ with state fixed effects and time fixed effects. Specifically, we used 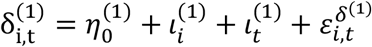 to estimate the vaccination rate for the first dose and 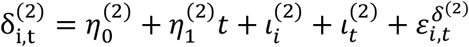 for the second dose. We predicted δ_i,t_ for each state in future periods based on the estimated constants 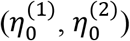, coefficient 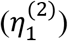, state fixed effects 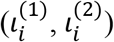, and the median of time fixed effects 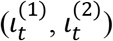

#### Vaccine efficacy (e)

According to previous studies, the Pfizer vaccine has an efficacy of 52.0% after the first dose, and the Moderna vaccine has an efficacy of 92.1% after the first dose^5^. Pfizer occupied a share of 47.75% of U.S. vaccine distribution during our sample period, and Moderna occupied the rest. We thus calculated the overall efficacy as the weighted average of both vaccines, at 73.0%.

#### Missing data imputation

Due to the lack of recovery data, we only know the cumulative infection rate rather than the current infection rate in the 22 states for which recovery data are missing. To produce quantitative results as accurately as possible, we used our SIR model to impute missing data for these 22 states. We first estimated vaccination coverage 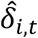 and the infection rate 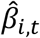 for these 22 states. Then, given the current infection rate at the initial period 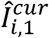, we calculated 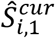 and generated the dynamics of cumulative infection rate 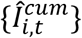 using

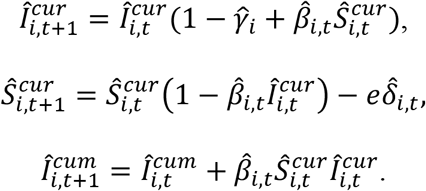

We then matched the model generated 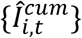 from the equations above for each of the 22 states with observed data for cumulative infection 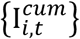 by minimizing the loss function below

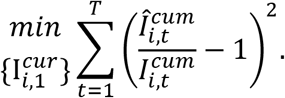

Supplementary Table 1 shows estimated region fixed effects. Supplementary Table 2 presents the estimated time fixed effects. Other model parameter input values can be found in Supplementary Table 3.

### 1.3 Model Fit

We examined how well our calibrated model fits the empirical data. The infection rates predicted by our model match the general trend in the U.S. and in most states quite well (see Supplementary Table 1 and Extended Data Fig. 7); the average correlation was 99.69% at the national level. Table 1 compares the fitness of the model results with the empirical data for each state. There were two exceptions, Kentucky (KY) and Maryland (MD), for which our model predictions were off-target by relatively large margins. However, this was due to the estimated removal rates (*γ*_*i*_) for these two states, which are outliers (Supplementary Fig. 1).

### 1.4 Model Predictions and Herd Immunity

The basic reproduction number, 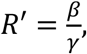 is the key measure used to assess the dynamics of the pandemic and to calculate the vaccination coverage to achieve herd immunity^6,7^. It is worthy of note that this formula only applies during the early stage of disease when the susceptible density approaches 1. However, at a later stage of the pandemic and with vaccines, a considerable share of the susceptible population has been vaccinated or has recovered, so the share of susceptible individuals can be significantly less than 1. According to the definition of the basic reproduction number, at period *t*, an infected person is expected to infect β_t_*S*_*t*_ people with an expected duration infection time of 1/γ. Therefore, the time-varying reproduction number is *R*′_*t*_= β_t_*S*_*t*_/*γ*. At the beginning of the pandemic, we have *t* = 0, *S*_*t*_ = 1 and *R*_0_ = β_0_/*γ*, which is consistent with the conventional definition. To assess whether the U.S. as a whole has acquired herd immunity, we use the “Third Statistics” approach; that is, the third-worst state’s reproduction number is used to form the national level “reproduction number”:

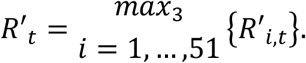

We used this measure to rule out the impact of outliers.^8^ As Supplementary Fig. 1 indicates, two states (Kentucky and Maryland) reported unreasonably low recovery numbers, which greatly biased our calculations of the reproduction number.

**Supplementary Fig. 1.**
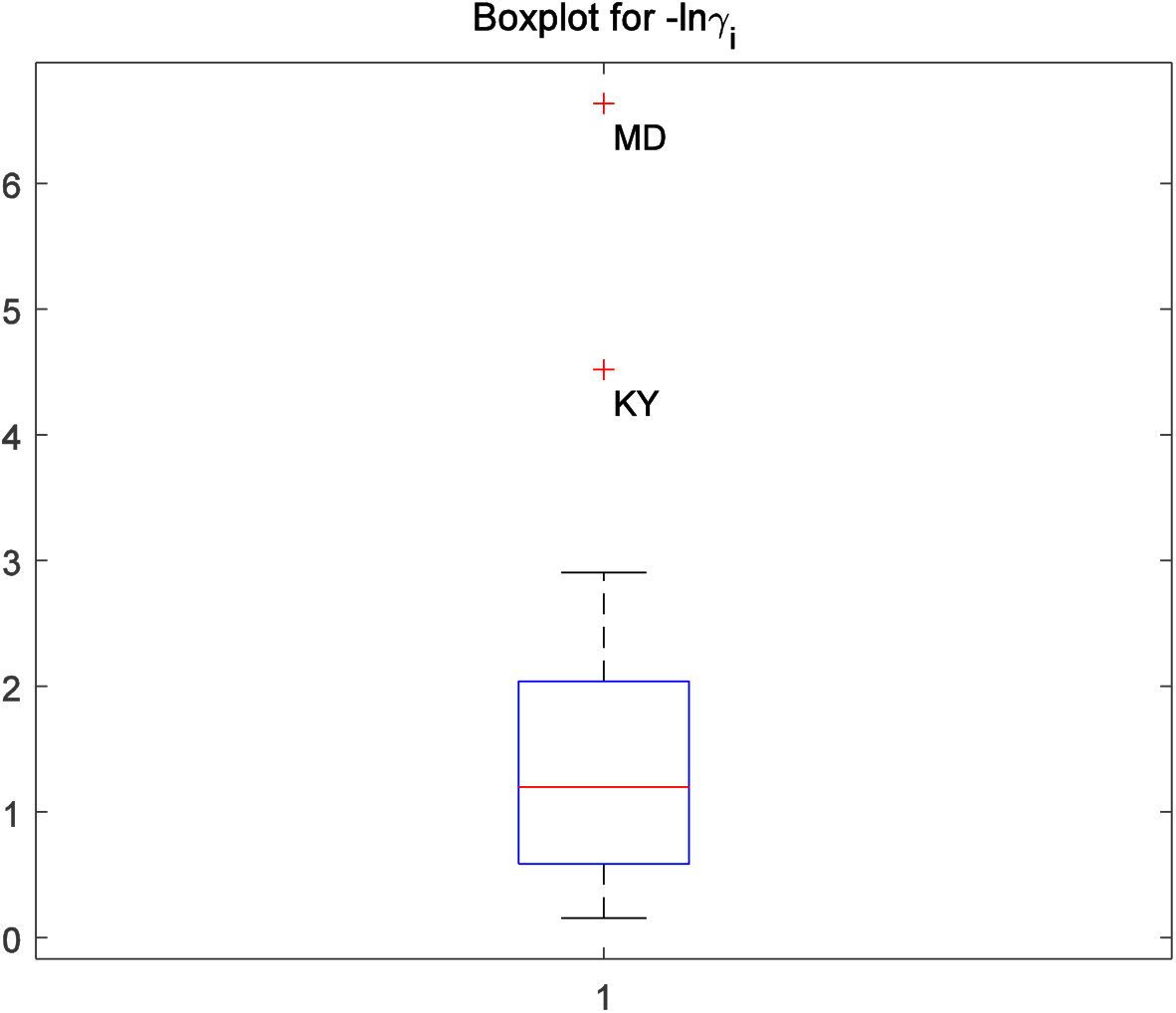
Boxplot for estimated state-level recovery rate. This figure plots the distribution of the estimated recovery rate for the 29 states with valid recovery data during our study period. According to the boxplot, Kentucky (KY) and Maryland (MD) appear as outliers.

**Supplementary Table 1. State fixed effects and model fitness across all 50 states and DC.** (in attached Excel file due to table size)

**Supplementary Table 2.** Time fixed effects across all 50 states and DC.

| Time | Estimated with First-Dose Data |  | Estimated with Second-Dose Data |  |
| --- | --- | --- | --- | --- |
|  | Infection Rate | Vaccination Rate | Infection Rate | Vaccination Rate |
| | Fixed Effect<br>$\rho_t$ (%) | Fixed Effect<br>$\iota_t^{(1)}$ (%) | Fixed Effect<br>$\rho_t$ (%) | Fixed Effect<br>$\iota_t^{(2)}$ (%) |
| Week 1 | 0.00 | N/A | 0.00 | N/A |
| Week 2 | 3.83 | N/A | 3.83 | N/A |
| Week 3 | 8.91 | N/A | 8.91 | N/A |
| Week 4 | 16.35 | N/A | 16.36 | N/A |
| Week 5 | 2.41 | N/A | 2.44 | N/A |
| Week 6 | -7.23 | N/A | -7.18 | N/A |
| Week 7 | -5.49 | N/A | -5.43 | N/A |
| Week 8 | -7.32 | N/A | -7.27 | N/A |
| Week 9 | -12.08 | N/A | -12.02 | N/A |
| Week 10 | -16.36 | N/A | -16.30 | N/A |
| Week 11 | -11.68 | N/A | -11.62 | N/A |
| Week 12 | -3.07 | N/A | -3.01 | N/A |
| Week 13 | -12.70 | N/A | -12.66 | N/A |
| Week 14 | -18.39 | 0.00 | -19.14 | 0.00 |
| Week 15 | -21.14 | -1.08 | -22.20 | -0.21 |
| Week 16 | -22.29 | -1.46 | -23.50 | -0.13 |
| Week 17 | -24.19 | -1.58 | -25.41 | -0.02 |
| Week 18 | -24.66 | -1.47 | -25.95 | 0.10 |
| Week 19 | -21.73 | -0.83 | -23.30 | 0.33 |
| Week 20 | -15.15 | -1.27 | -17.04 | 0.23 |
| Week 21 | N/A | -0.63 | N/A | 0.00 |

**Supplementary Table 3.** Other model parameter values.

|  | **Notations** | **Values** |
| --- | --- | --- |
| **Estimations with**<br>**First Dose Data** | Constant of Infection Rate Estimation ($\theta_0$) | 0.391 |
| | Elasticity of Policy on Infection Rate Estimation ($\theta_1$) | 0.00529 |
| | Constant of Vaccination Rate Estimation ($\eta_0^{(1)}$) | 0.0285 |
| **Estimations with**<br>**Second Dose Data** | Constant of Infection Rate Estimation ($\theta_0$) | 0.397 |
| | Elasticity of Policy on Infection Rate Estimation ($\theta_1$) | -0.00542 |
| | Constant of Vaccination Rate Estimation ($\eta_0^{(2)}$) | 0.00142 |
| | Trend of Vaccination Rate Estimation ($\eta_1^{(2)}$) | 0.00185 |

## Acknowledgments

We thank K.E. Warner and S. Mennemeyer for their feedback. Funding: H.H. is supported by the startup grant from the City University of Hong Kong (grant no. 7200689).

## Author contributions

All authors designed the analyses, interpreted the results, and designed the figures, and are listed alphabetically. X.C., H.H., R.S., and J.Z. contribute equally to the paper. H.H. and R.S. collected the data. J.Z. conducted the reduced-form empirical analysis. X.C. conducted the analysis with the SIR model. H.H., J.J., and R.S. wrote the paper.

## Data and Code Availability

**Extended Data Fig. 1.**
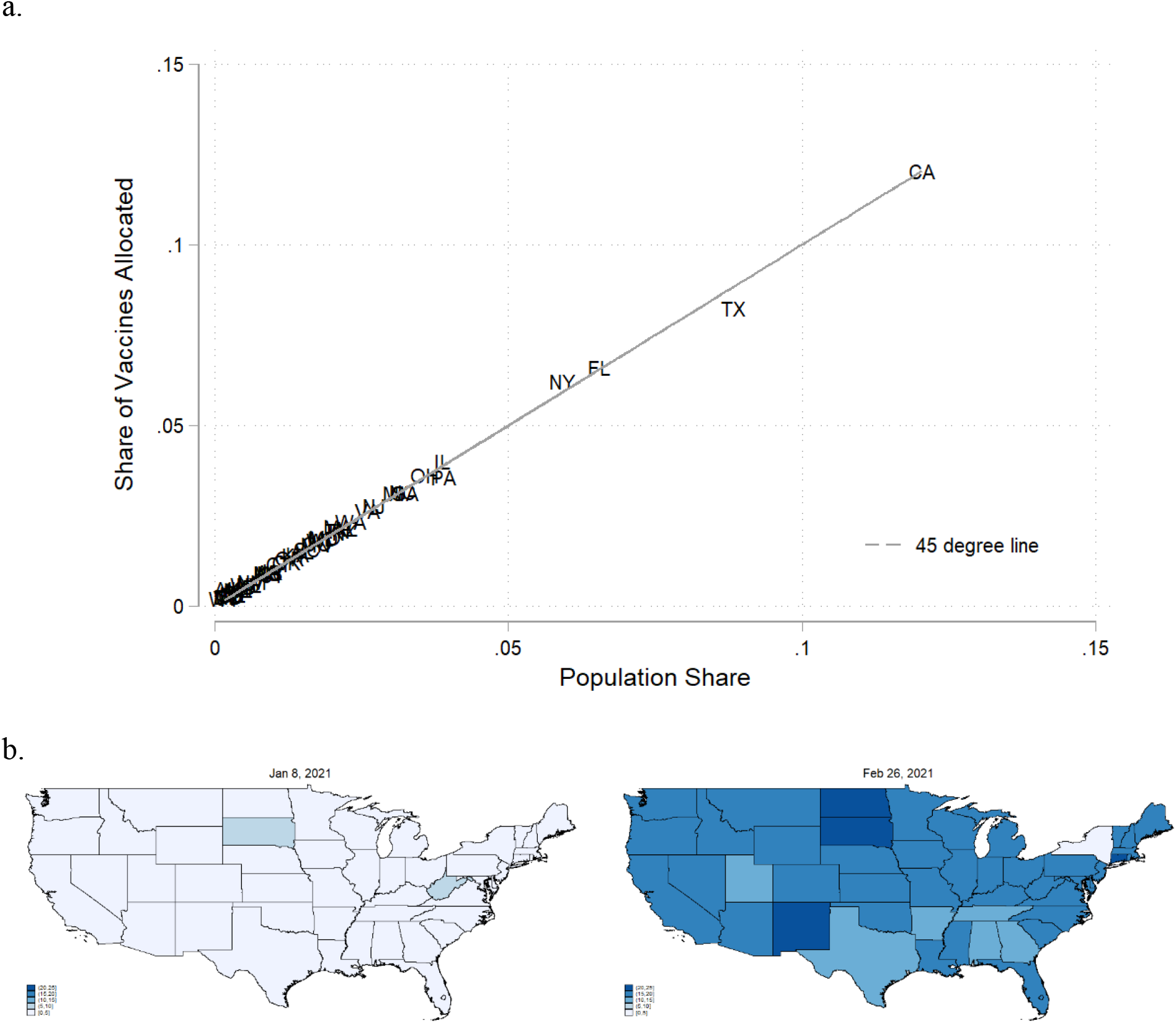
COVID-19 vaccination in all 50 U.S. states and DC. **a**, Share of vaccines allocated versus population share. **b**, Heat map of vaccines administered by states over time. The darker the color, the more doses of vaccines administered per 100 people.

**Extended Data Fig. 2.**
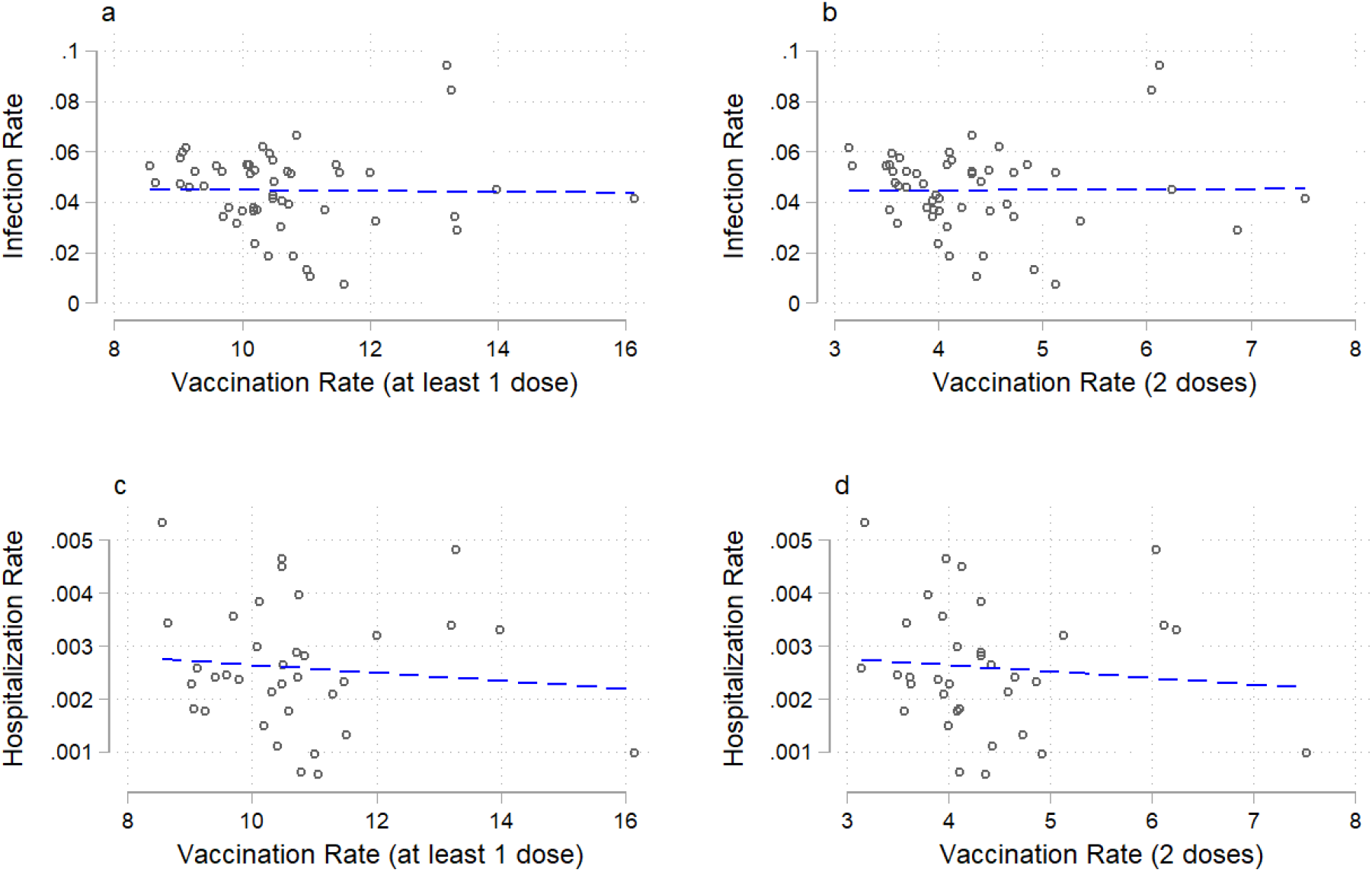
COVID-19 infections (average total infection and hospitalization rates) before vaccination and average vaccination rate. **a**, Association between the total infection rate before vaccination and at least 1 dose of vaccination (coefficient = 0.0002, R^2^ = 0.0%). **b**, Association between the total infection rate before vaccination and 2 doses of vaccination (coefficient = 0.0002, R^2^ = 0.0%). **c**, Association between the total hospitalization rate before vaccination and at least 1 dose of vaccination (coefficient = 0.0000, R^2^ = 1.0%). **d**, Association between the total hospitalization rate before vaccination and 2 doses of vaccination (coefficient = 0.0001, R^2^ = 0.9%).

**Extended Data Fig. 3.**
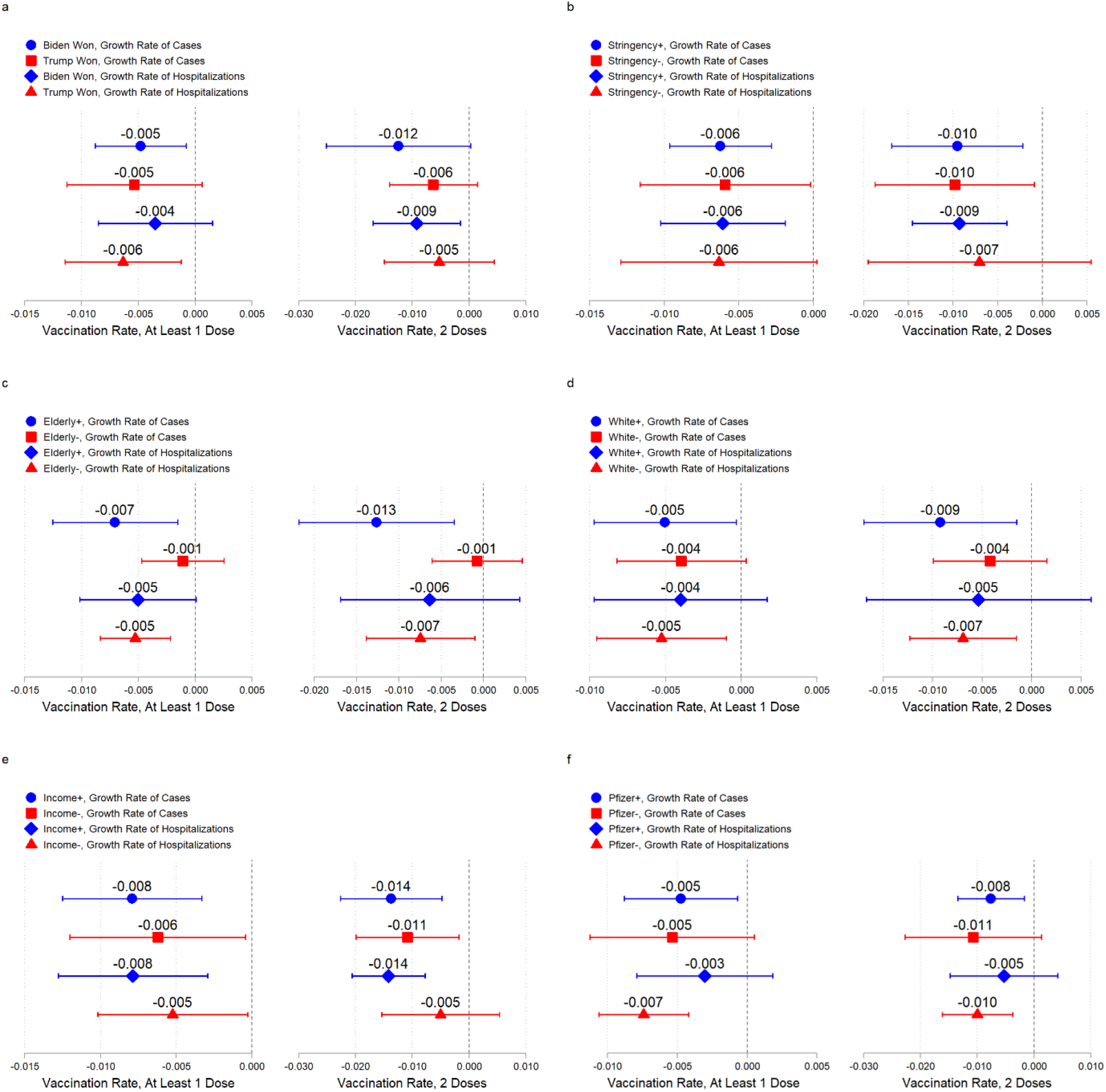
Heterogeneity tests on the effect of vaccination across various state characteristics. Blue markers are the estimated effects of at least 1 dose of vaccine, and red markers are the estimated effects of 2 doses of vaccine. **a**, Effect of vaccination in states where the 2020 presidential election was won by Joe Biden versus Donald Trump. **b**, Effect of vaccination in states with non-pharmaceutical interventions more stringent than the national median (+) versus less stringent than the median (-). **c**, Effect of vaccination in states with the proportion of the elderly population (65+) greater than the national median (+) versus less than the median (-). **d**, Effect of vaccination in states with the proportion of the white population greater than the national median (+) versus less than the median (-). **e**, Effect of vaccination in states with per capita income greater than the national median (+) versus less than the median (-). **f**, Effect of vaccination in states with the share of Pfizer vaccine greater than the national median (+) versus less than the median (-).

**Extended Data Fig. 4.**
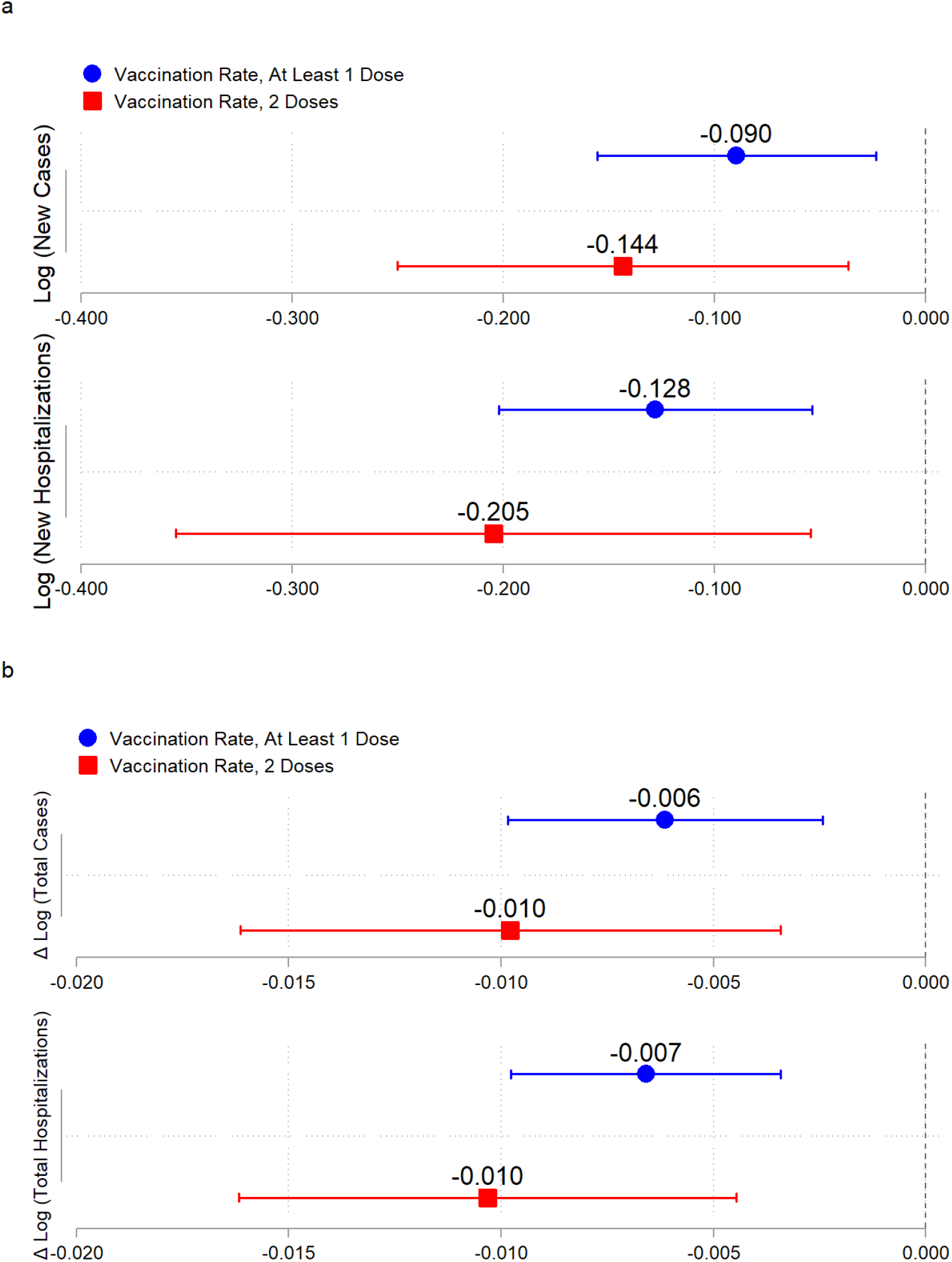
Estimated effects of vaccination on the COVID-19 pandemic with alternative outcome measures. Blue markers are the estimated effects of at least 1 dose of vaccine, and red markers are the estimated effects of 2 doses of vaccine. **a**, Estimated effects of vaccination on logarithms of news cases and hospitalizations. **b**, Estimated effects of vaccination on changes in logarithms of total cases and hospitalizations.

**Extended Data Fig. 5.**
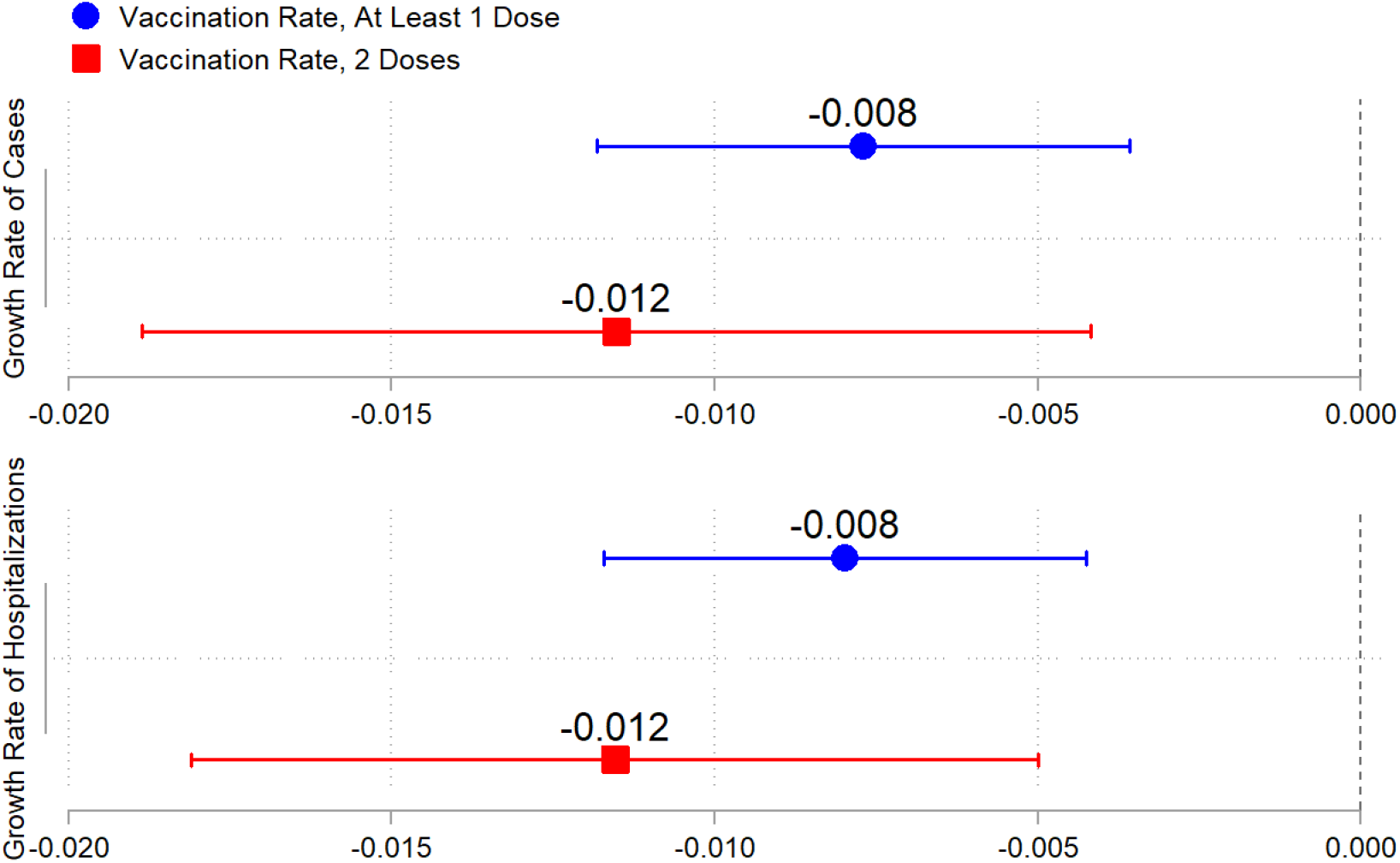
Estimated effects of vaccination on the COVID-19 pandemic with imputed missing data on vaccination between 21 December 2020 and 10 January 2021. Blue markers are the estimated effects of at least 1 dose of vaccine, and red markers are the estimated effects of 2 doses of vaccine.

**Extended Data Fig. 6.**
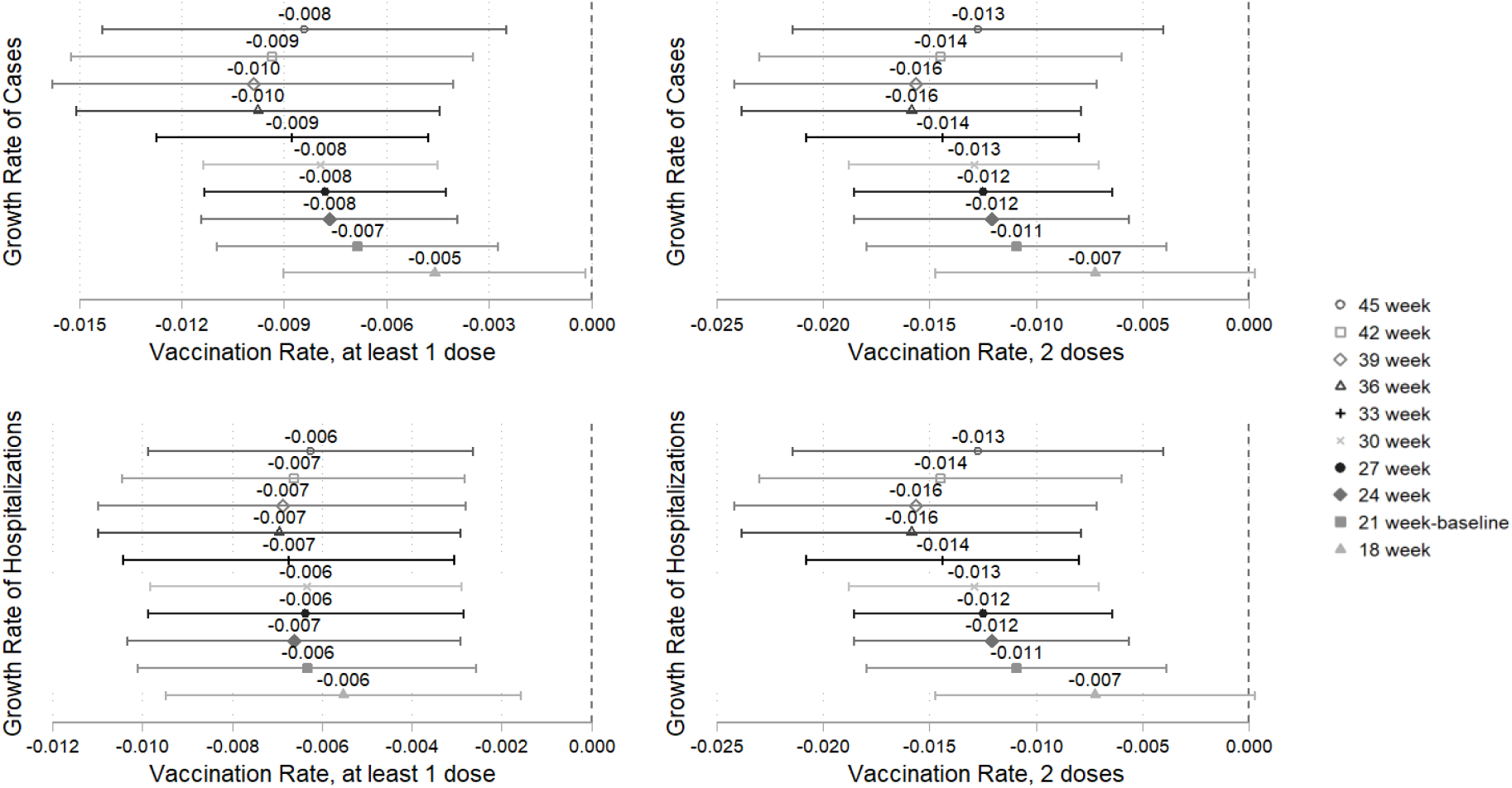
Estimated effects of vaccination on the COVID-19 pandemic with different sample periods. Our 21-week baseline period is from 12 October 2020 to 7 March 2021. 18-week period is from 2 November 2020 to 7 March 2021; 24-week from 21 September 2020 to 7 March 2021; 27-week from 31 August 2020 to 7 March 2021; 30-week from 10 August 2020 to 7 March 2021; 33-week from 20 July 2020 to 7 March 2021; 36-week from 29 June 2020 to 7 March 2021; 39 week from 8 June 2020 to 7 March 2021; 42-week from 18 May 2020 to 7 March 2021; and 45-week from 27 April 2020 to 7 March 2021.

**Extended Data Fig. 7.**
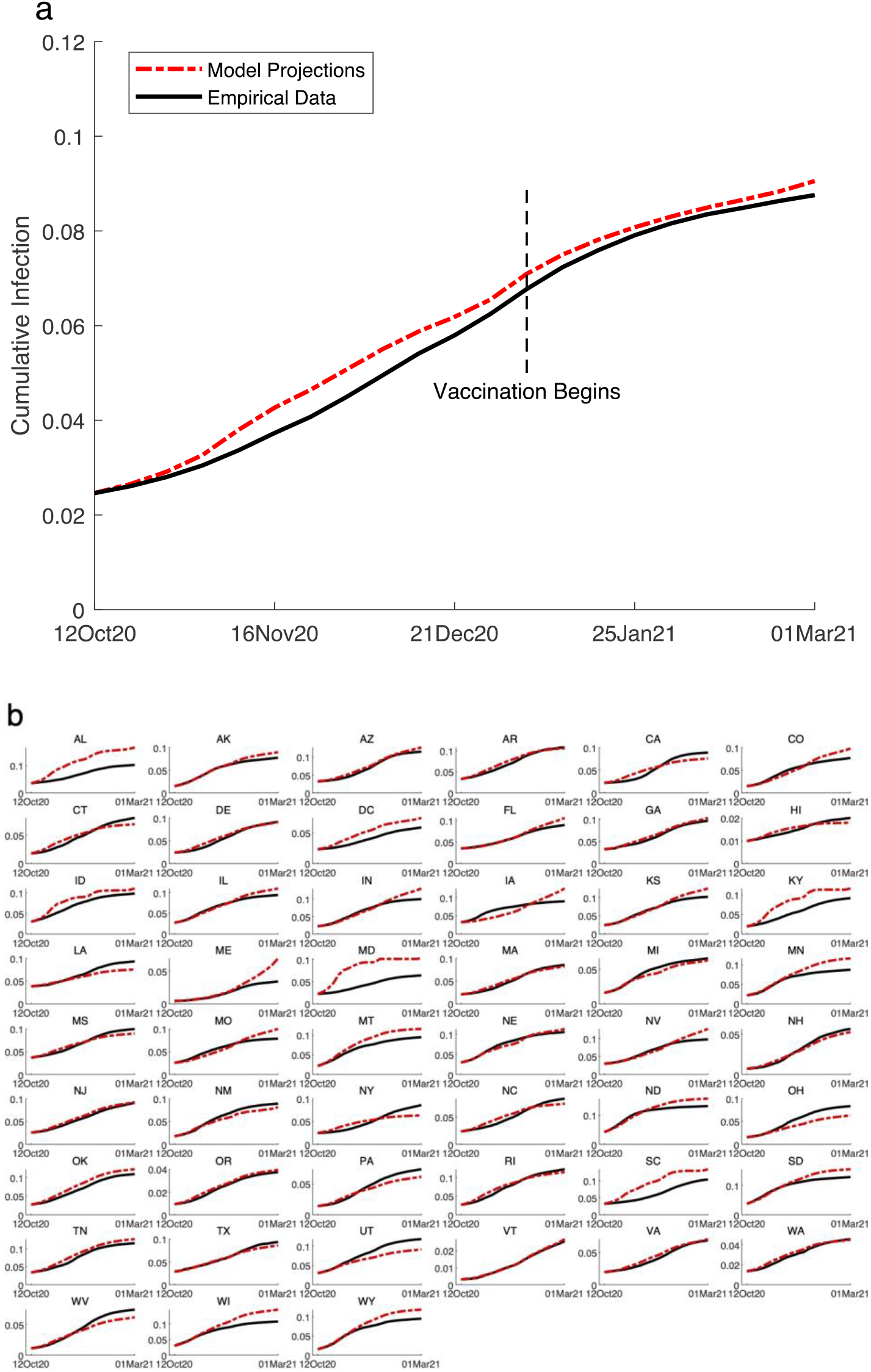
Development of cumulative infection rate during our study period (12 October 2020 to 7 March 2021). Red curves are model projections, and black curves are empirical data. **a**, National cumulative infection rate, model projections versus empirical data. Our model projections are 99.69% correlated with empirical data. **b**, Development of cumulative infection rate across all 50 U.S. states and DC, model projections versus empirical data. Our model estimations reached a median correlation of 99.04% with empirical data, with a minimum of 86.37% in Maryland and a maximum of 99.95% in Vermont.

**Extended Data Table 1.** Summary statistics.

| **Variable** | **N** | **Mean** | **SE<sup>a</sup>** | **Median** | **(Min, Max)** |
| --- | --- | --- | --- | --- | --- |
| Growth of total cases | 1071 | 0.07 | 0.05 | 0.06 | (0, 0.3) |
| New cases | 1071 | 19583.4 | 28962.1 | 10544 | (72, 302690) |
| Growth of total hospitalizations | 735 | 0.05 | 0.04 | 0.04 | (0, 0.5) |
| New hospitalizations | 735 | 629.4 | 798.8 | 417 | (5, 13127) |
| Stringency index | 1071 | 46.3 | 13.4 | 46.3 | (7.4, 75.9) |
| People vaccinated per hundred<sup>b</sup> | 1071 | 4.1 | 6.1 | 0 | (0, 24.7) |
| People fully vaccinated per hundred<sup>c</sup> | 1071 | 1.7 | 3.0 | 0 | (0, 15.6) |
| Susceptible / population | 1071 | 94.1 | 2.9 | 94.3 | (86.8, 99.7) |
| Testing / population | 1071 | 80.1 | 41.9 | 71.6 | (20.0, 295.4) |
| Rally (dummy variable) | 1071 | 0.03 | 0.2 | 0 | (0, 1) |
| Number of protests | 1071 | 0.06 | 0.4 | 0 | (0, 7) |
| Snow depth | 1071 | 19.5 | 34.0 | 4.5 | (0, 243.3) |
| Deviation from mean temperature<sup>d</sup> | 1071 | -0.9 | 0.7 | -1.0 | (-3.1, 0.7) |

Notes.
<sup>a</sup> Standard error.
<sup>b</sup> People who received at least 1 dose of the vaccine.
<sup>c</sup> People who received 2 doses of the vaccine.
<sup>d</sup> Deviation from mean temperature = (temperature at state i week t – mean temp of state i during 12 Oct 2020 to 7 Mar 2021) / standard error of temperature of state i.

**Extended Data Table 2.** Baseline regression results.

| **Variables** | **Growth rate of cases** |  | **Growth rate of hospitalizations** |  |
| --- | --- | --- | --- | --- |
|  | **(N = 1,071)** |  | **(N = 735)** |  |
| People vaccinated per hundred<sup>a</sup> | -0.07***<br>(0.002) |  | -0.007***<br>(0.002) |  |
| People fully vaccinated per hundred<sup>b</sup> |  | -0.011***<br>(0.004) |  | -0.011***<br>(0.003) |
| Stringency index | -0.000<br>(0.001) | -0.000<br>(0.001) | 0.001<br>(0.000) | 0.000<br>(0.000) |
| Susceptible rate | 0.021***<br>(0.005) | 0.021***<br>(0.005) | 0.010***<br>(0.003) | 0.010***<br>(0.003) |
| Test rate | 0.000<br>(0.000) | 0.000<br>(0.000) | 0.000<br>(0.000) | 0.000<br>(0.000) |
| Rally | -0.004<br>(0.007) | -0.004<br>(0.007) | -0.005<br>(0.008) | -0.005<br>(0.008) |
| Number of protests | -0.001<br>(0.003) | -0.000<br>(0.003) | 0.001<br>(0.006) | 0.001<br>(0.006) |
| Log (snow depth) | 0.001<br>(0.002) | 0.001<br>(0.002) | 0.000<br>(0.001) | 0.001<br>(0.001) |
| Deviation from mean temperature<sup>c</sup> | -0.003<br>(0.006) | -0.003<br>(0.006) | -0.008<br>(0.006) | -0.008<br>(0.006) |
| R-squared | 71.4% | 71.5% | 48.2% | 48.1% |

Notes.
Standard errors (in parentheses) are two-way clustered at state and week level.
Significance levels: \*\*\* $p < 0.01$ , \*\* $p < 0.05$ , \* $p < 0.1$ .
<sup>a</sup> People who received at least 1 dose of the vaccine.
<sup>b</sup> People who received 2 doses of the vaccine.
<sup>c</sup> 21 observations of DC temperature were missing, estimated using average temperature from neighboring states of Virginia and Maryland. 8 observations of Delaware temperature were missing, estimated using average temperature from the neighboring states of New Jersey and Maryland.

**Extended Data Table 3.** Baseline regression results with data of daily frequency.

| Variables | Growth rate of cases<br>(N = 7,497) |  | Growth rate of hospitalizations<br>(N = 5,145) |  |
| --- | --- | --- | --- | --- |
| People vaccinated per hundred<sup>a</sup> | -0.0009***<br>(0.0002) |  | -0.0009***<br>(0.0002) |  |
| People fully vaccinated per hundred<sup>b</sup> |  | -0.0015***<br>(0.0003) |  | -0.0015***<br>(0.0003) |
| Stringency index | 0.0001<br>(0.0001) | 0.0001<br>(0.0001) | 0.0001*<br>(0.0001) | 0.0001<br>(0.0001) |
| Susceptible rate | 0.0027***<br>(0.0005) | 0.0027***<br>(0.0005) | 0.0012***<br>(0.0003) | 0.0013***<br>(0.0003) |
| Test rate | 0.0000<br>(0.0000) | 0.0000<br>(0.0000) | 0.0000<br>(0.0000) | 0.0000<br>(0.0000) |
| Rally | -0.0002<br>(0.0013) | -0.0002<br>(0.0013) | -0.0007<br>(0.0016) | -0.0007<br>(0.0016) |
| Number of protests | -0.0000<br>(0.0009) | 0.0000<br>(0.0010) | -0.0007<br>(0.0008) | -0.0007<br>(0.0008) |
| Log (snow depth) | -0.0002<br>(0.0002) | -0.0002<br>(0.0002) | -0.0000<br>(0.0002) | 0.0000<br>(0.0002) |
| Deviation from mean temperature<sup>c</sup> | -0.0007*<br>(0.0004) | -0.0007*<br>(0.0004) | -0.0004<br>(0.0004) | -0.0004<br>(0.0004) |
| R-squared | 51.7% | 51.8% | 16.4% | 16.4% |

Notes.
Standard errors (in parentheses) are two-way clustered at state and week level.
Significance levels: \*\*\* p<0.01, \*\* p<0.05, \* p<0.1.
<sup>a</sup> People who received at least 1 dose of the vaccine.
<sup>b</sup> People who received 2 doses of the vaccine.
<sup>c</sup> 21 observations of DC temperature were missing, estimated using average temperature from neighboring states of Virginia and Maryland. 8 observations of Delaware temperature were missing, estimated using average temperature from the neighboring states of New Jersey and Maryland.

## Footnotes

1 The states with valid recovery data are AL, AR, DC, ID, KY, LA, ME, MD, MA, MI, MN, MS, MT, NE, NH, NM, ND, OH, OK, PA, SC, SD, TN, TX, UT, VT, WV, WI, and WY. Although IA also reported recovery, the number was higher than the cumulative number of infections. We therefore excluded IA as well.

2 Hsiang (2020)

3 We adopted the median instead of the mean to dampen the influence of outliers, similar to Hsiang (2020).

4 See footnote 1 for details, in which we list all states with complete recovery data.

5 Creech et al, 2021

6 Sun, 2010

7 Sun & Shi, 2011

8 Eaton & Kortum, 2002

